# Analytical Solution of a New *SEIR* Model Based on Latent Period-Infectious Period Chronological Order

**DOI:** 10.1101/2021.12.14.21267812

**Authors:** Xiaoping Liu

**Affiliations:** Department of Medicine, Department of Neuroscience, Rockefeller Neuroscience Institute, West Virginia University Health Science Center, Morgantown, WV 26506 USA

## Abstract

The Susceptible-Infectious-Recovered (*SIR*) and *SIR* derived epidemic models have been commonly used to analyze the spread of infectious diseases. The underlying assumption in these models, such as Susceptible-Exposed-Infectious-Recovered (*SEIR*) model, is that the change in variables *E, I* or *R* at time *t* is dependent on a fraction of *E* and *I* at time *t*. This means that after exposed on a day, this individual may become contagious or even recover on the same day. However, the real situation is different: an exposed individual will become infectious after a latent period (*l*) and then recover after an infectious period (*i*). In this study, we proposed a new *SEIR* model based on the latent period-infectious period chronological order (Liu X., Results Phys. 2021; 20:103712). An analytical solution to equations of this new *SEIR* model was derived. From this new *SEIR* model, we obtained a propagated curve of infectious cases under conditions *l*>*i*. Similar propagated epidemic curves were reported in literature. However, the conventional *SEIR* model failed to simulate the propagated epidemic curves under the same conditions. For *l*<*i*, the new *SEIR* models generated bell-shaped curves for infectious cases, and the curve is near symmetrical to the vertical line passing the curve peak. This characteristic can be found in many epidemic curves of daily COVID-19 cases reported from different countries. However, the curve generated from the conventional *SEIR* model is a right-skewed bell-shaped curve. An example for applying the analytical solution of the new *SEIR* model equations to simulate the reported daily COVID-19 cases was also given in this paper.

## INTRODUCTION

*SIR* (Susceptible-Infectious-Recovered) and *SIR* derived epidemic mathematical models (such as *SEIR* model; Susceptible-Exposed-Infectious-Recovered) are commonly used in the analysis of transmission of infectious diseases^1-6^. These models have been playing an important role in formulating the proper social interventions to slow down the spread of COVID-19^7-15^. In the conventional *SEIR* model, the relationship between the four variables *S, E, I* and *R* is defined by the following differential equations^2^:

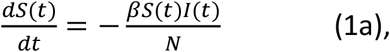

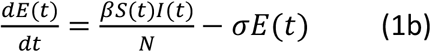

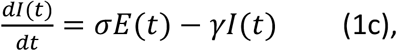

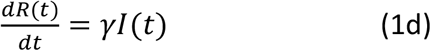

In Eqn. (1), *N* is the total population; and the coefficients *β, σ* and *γ* are constants. One of the underlying assumptions for the above equations are: The change in *E, I* or *R* on a day is determined by a fraction of *E* and *I* (*σE* and *γI* in Eqn. (1)) on that day. This means that if a susceptible individual is exposed to an infectious source on a day, the exposed individual will be able to become an infectious individual immediately together with all others who were exposed in previous days at constant fraction coefficients *σ* and *γ* without chronological order. However, the reality is different: after a susceptible individual is exposed to infectious individuals, the newly exposed individual cannot become an infectious individual until the exposed individual passes a latent period. Likewise, an individual in the infectious period can only recover after an infectious period. Thus, the underlying assumptions of the conventional *SIR* or *SIR*-derived models are not optimized for describing the transmission process of infectious diseases. A latent period-infectious period chronological order is an inherent relationship that exists among the 4 variables, and a new epidemic model based on the latent period-infectious period chronological order is desirable.

In this study, we will develop a new *SEIR* model based on the latent period-infectious period chronological order^16,17^. The analytic solution of the related model equations will be presented. We will demonstrate the similarities and differences between the new *SEIR* model and the conventional *SEIR* model, and show the improvement made by the new *SEIR* model in simulations of the epidemic curves.

## THEORY

The new *SEIR* model is based on the following three assumptions:

1. During the spread of an infectious disease, the decreasing rate of the number of susceptible individuals (*S*) is proportional to the number of infectious individuals (*I*) and proportional to the ratio of *S* to the population *N* (*S/N*). This assumption is the same as the one described in the classic *SIR* model.
2. After a susceptible individual is exposed to an infectious source, the individual will become infectious after a latent period (*l* days in average) and then recover or die after an infectious period (*i* days in average). The total time length of the latent period and the infectious period is *c*, or *c*=*l*+*i*.
3. Only a fraction *α* of infected individuals who are in the infectious period (*I*_*n*_) are confirmed and reported as daily new cases (*y*_*n*_).

We can derive the following recursive equation (2a) from Assumption (1), write the following recursive equations (2b)-(2d) based on Assumption (1) and (2), and give the following equation (2e) based on Assumption (3).

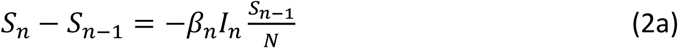

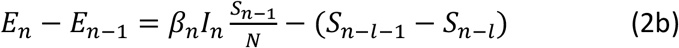

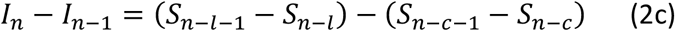

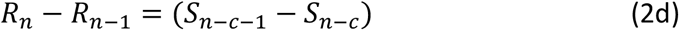

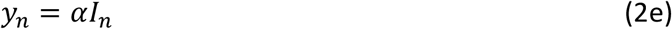

We further assume:

a. For all *n*<0 before the infectious disease starts to spread, *E*_*n*_*=I*_*n*_*=R*_*n*_*=*0 and *S*_*n*_=*N*, where *N* is the population.
b. At *n*=0, when the first person was infected by the infectious disease studied, *S*_0_=*N*-1, *E*_0_=1, and *I*_0_=*R*_0_=0.

Thus, we can obtain:

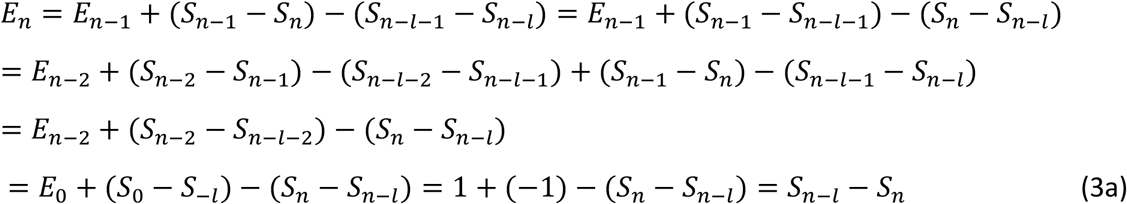

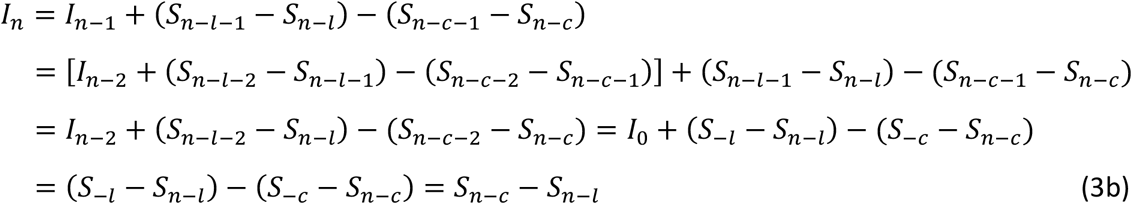

From Eqn. (2d) we have:

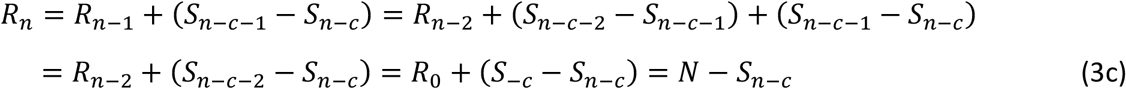

Substitution of Eqn. (3b) into Eqn. (2a) gives:

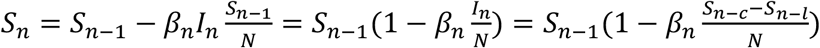

Thus, the final solution of *S*_*n*_ is the following formula:

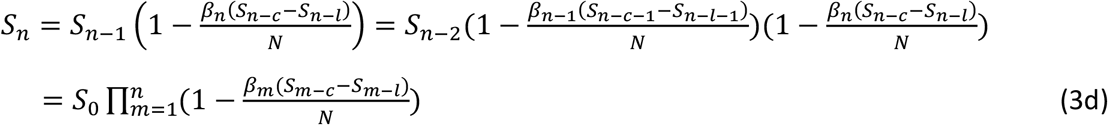

Finally, we obtain the following analytical solutions of variables *S*_*n*_, *E*_*n*_, *I*_*n*_ and *R*_*n*_ from Eqns. (3a)-(3d):

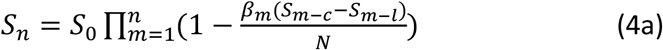

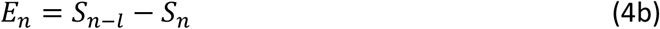

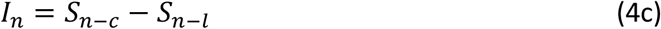

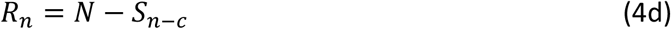

From Eqn. (4a), we can calculate the value of *S*_*n*_ on any day *n* if *l* and *c* are known and *β*_*n*_ is a constant. Then, values of *E*_*n*_, *I*_*n*_ and *R*_*n*_ can be calculated from Eqns. (4b)-(4d) after *S*_*n*_ are determined. If *β*_*n*_ varies with time, *β*_*n*_ on any day *n* can be determined by fitting *αI*_*n*_ to 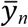 based on Eqn. (2e) assuming that *α* has been determined. In this case, we need both Eqn. (4a) and Eqn. (2e) to determine *S*_*n*_ and *β*_*n*_, and then find *E*_*n*_, *I*_*n*_ and *R*_*n*_ from Eqns. (4b)-(4d) after *β*_*n*_ is determined.

## RESULTS

To calculate *S*_*n*_, *E*_*n*_, *I*_*n*_ and *R*_*n*_ in this new *SEIR* model, we can input Eqns. (4a)-(4d) into Excel. Thus, for a given pair of parameters *l* and *c* (*c=l+i*), we can calculate *S*_*n*_, *E*_*n*_, *I*_*n*_ and *R*_*n*_ in the Excel program assuming *β*_*n*_=1. We first compared the number of infectious individuals *I*_*n*_ calculated from the new *SEIR* model with the one calculated from the conventional *SEIR* model assuming *l*>*i* (Figure 1) and *l*<*i* (Figure 2). It can been seen that the new *SEIR* model generated a propagated epidemic curve for the number of infectious individuals as *l*>*i* by assuming *l*=1/*σ* =8, *i*=1/*γ* =2 and *N*=3.3×10^8^ (Figure 1A). However, under the same conditions (*l*=1/*σ* =8, *i*=1/*γ* =2 and *N*=3.3×10^8^), the conventional *SEIR* model generated an epidemic curve, which increased in a near-exponential form, for the number of infectious individuals (Figure 1B). We then compared the variables *S, E, I* and *R* calculated from the new *SEIR* model with those calculated from the conventional *SEIR* model as *l*<*i* by assuming *l*=1/*σ* =2, *i*=1/*γ* =8 (Figure 2). Under these conditions, both the new and the conventional *SEIR* models generated sigmoidal curves for the variables *S* and *R*, and bell-shaped curves for the variables *E* and *I*. However, the bell-shaped curves for the variable *I* that was generated from the new *SEIR* model were near symmetrical about the vertical line passing the peak (the red curve in Figure 2A); in contrast, the conventional *SEIR* model generated curve for the variable *I* was in a right-skewed bell shape (the red curve in Figure 2B). In Figure 3, we demonstrated two typical epidemic curves recorded in the real world. The one in Figure 3A shows the daily measles cases (an example of propagated epidemic curves) reported in Aberdeen, South Dakota, USA from October 15, 1970 to January 16, 1971^18^. The other one in Figure 3B shows the daily COVID-19 cases (an example of bell-shaped epidemic curves) reported in South Africa from April 15, 2020 to April 30, 2021^19,20^. Finally, we applied Eqn. (4) and Eqn. (2e) to simulate the number of daily new COVID-19 cases in the United States (Figure 4). In the simulation, we let *l*=4, *i*=10 ^17^ and *α*=0.01453^21^. The time-dependent *β*_*n*_ was regulated for best fitting the calculated number of daily COVID-19 cases *αI*_*n*_ (blue line) to the reported 7-days average number of daily COVID-19 cases 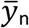 (red dots). In this way, we determined all *β*_*n*_ from 2/1/2020 to 1/6/2021. The value of the last *β*_*n*_ determined on January 6, 2021 is 0.17/day. Assuming that *β*_*n*_, is a constant after January 6, 2021, we calculated the number of daily COVID-19 cases in the United States. The calculated number of daily COVID-19 cases is consistent with the reported number of daily COVID-19 cases from early 2020 up to January 22, 2021.

**Figure 1.**
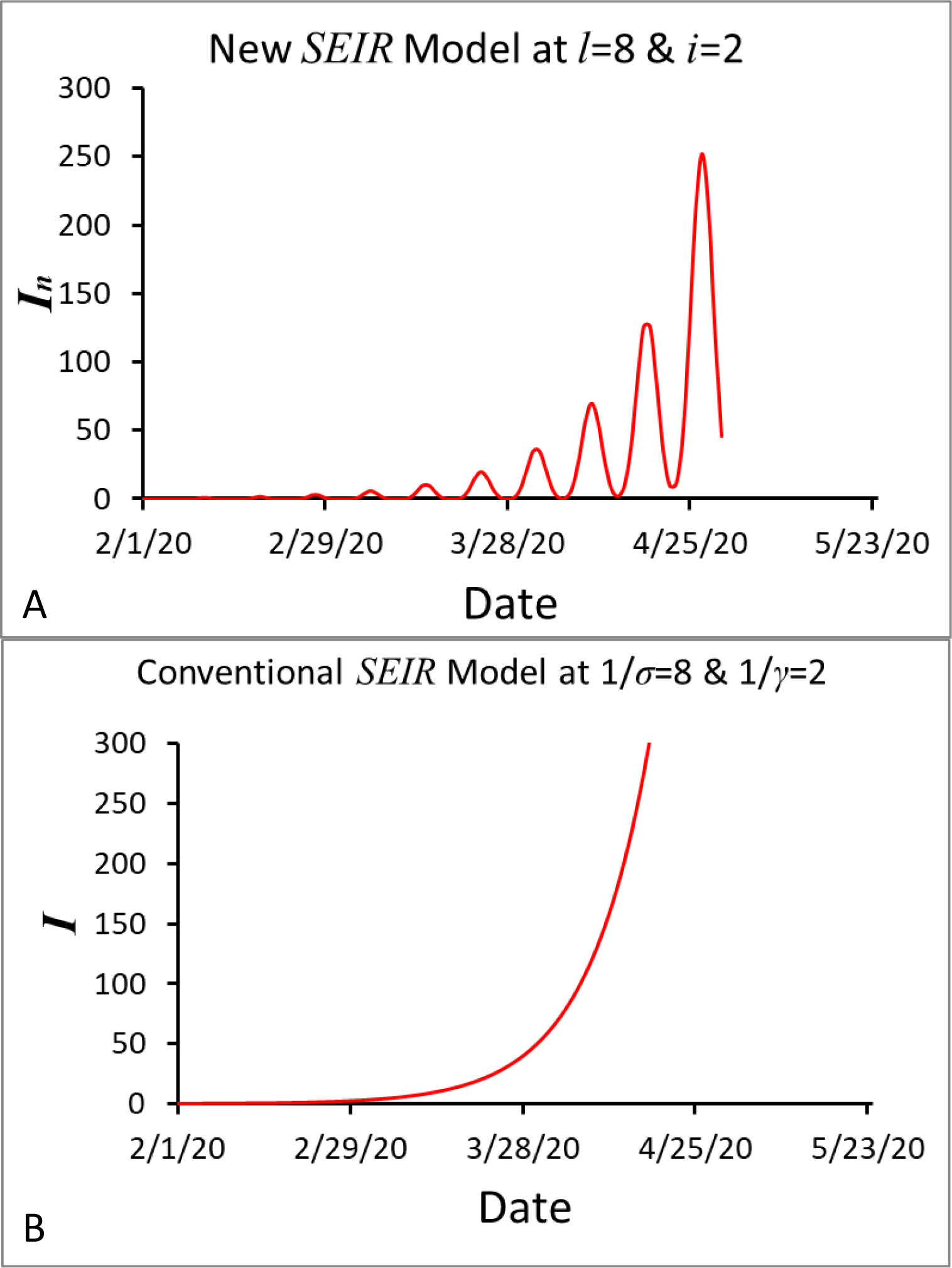
Comparison of the number of infectious individuals calculated from the new *SEIR* model (A) with the number of infectious individuals calculated from the conventional *SEIR* model (B) assuming that the latent period *l*=1/*σ*=8 days and the infectious period *i*=1/*γ*=2 days.

**Figure 2.**
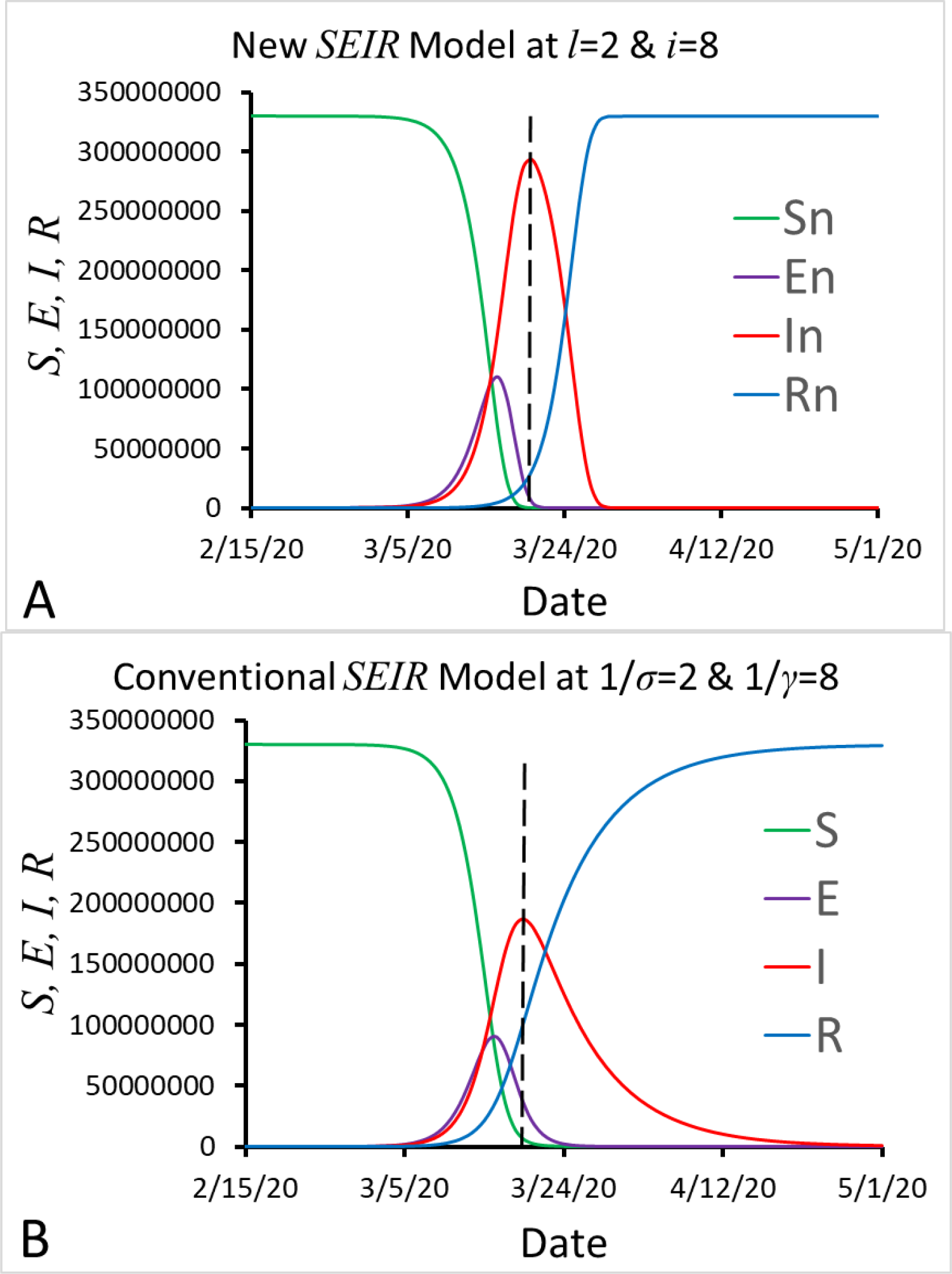
Comparison of *S, E, I* and *R* calculated from the new *SEIR* model (A) with *S, E, I* and *R* calculated from the conventional *SEIR* model (B) assuming that the latent period *l*=1/*σ*=2 days and the infectious period *i*=1/*γ*=8 days.

**Figure 3.**
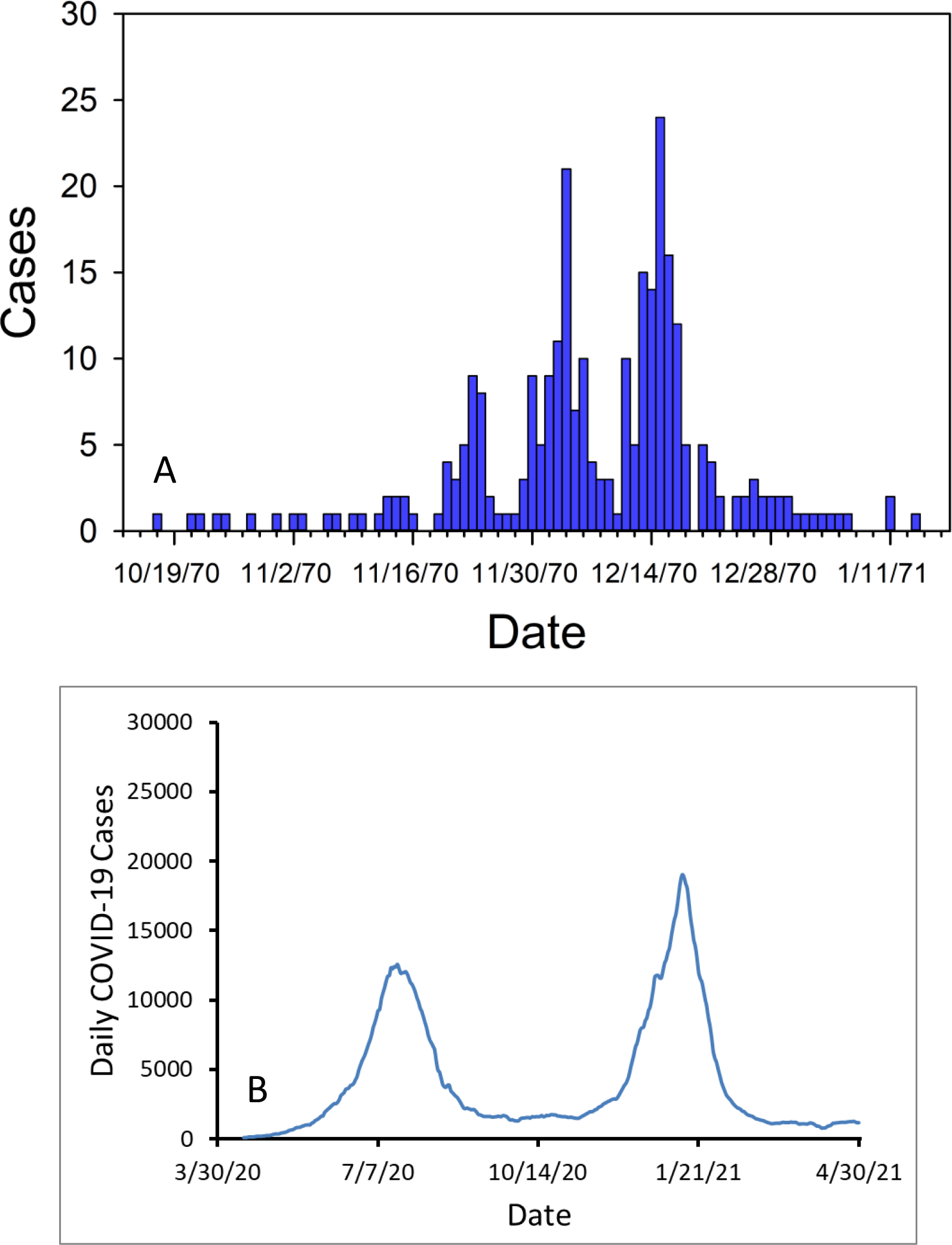
Daily measles cases (an example of propagated epidemic curves) reported in Aberdeen, South Dakota, USA from October 15, 1970 to January 16, 1971 (A) and daily COVID-19 cases reported in South Africa from April 15, 2020 to April 30, 2021 (B).

**Figure 4.**
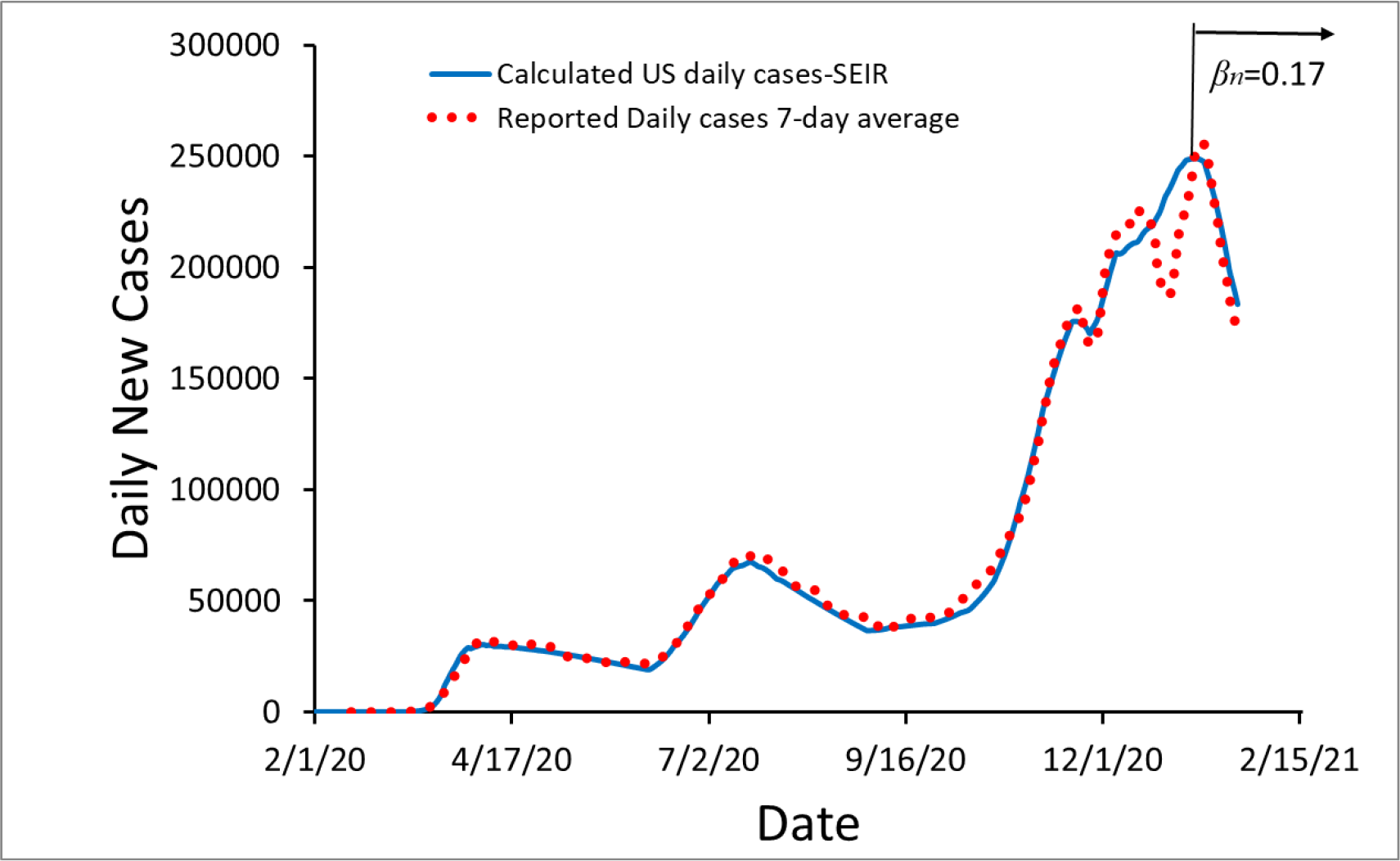
The number of daily new COVID-19 cases reported from 2/15/2020 to 1/22/2021 in the United States (red dots) and the number of daily new COVID-19 cases calculated from the new *SEIR* model in the same period (blue line).

## DISCUSSION

From Figure 1 we can see that if *l*>*i*, the curves representing the number of infectious individuals (*I*) calculated from the two models are largely different. The new *SEIR* model can generate a propagated epidemic curve that is similar to the reported propagated epidemic curve in Figure 3A. In contrast, the conventional *SEIR* model (Figure 1B) does not simulate a propagated epidemic curve even if the average latent period (1/*σ*=8) is greater than the average infectious period (1/*γ*=2). Why the new *SEIR* model can but the conventional *SEIR* model can’t simulate the propagated curve when latent period is longer than the infectious period? If we assume that the latent period *l*=10 days, the infectious period *i*=2 days, and that 10 infected people are all currently in the infectious period, and they cause 20 new people to become infected during the 2 days of infectious period, then we will only see a peak with 10 cases of infectious people in the current infectious period. The reason is that the newly infected 20 people are still in the latent period, and we can only see the next peak with the 20 new cases after the 10 days of latent period. Keeping transmission in this way, we will observe a propagated epidemic curve. This result indicates that the chronological order of latent period-infectious period should be applied to epidemic models for assuring the correct relationship among the model variables, and that the underlying assumptions of the conventional *SIR* or *SIR*-derived models are not optimized for describing the transmission process of infectious diseases.

From Figure 2 we can see that the curves *S, E, I* and *R* calculated from the new *SEIR* model (Figure 2A) are similar to those calculated from the conventional *SEIR* model (Figure 2B). However, both sides of the peak of curve *I*_*n*_ calculated from the new *SEIR* model (the red line in Figure 2A) are nearly symmetrical to the vertical dashed line passing through the peak of the curve *I*_*n*_. In contrast, both sides of the peak of curve *I* calculated from the conventional model (the red line in Figure 2B) do not have these symmetrical characteristics. It can be seen from Figure 3B that the curve shape of the daily new cases reported in South Africa (Figure 3B) is closer to the curve *I*_*n*_ calculated from the new *SEIR* model (Figure 2A) rather than the curve *I* calculated from the conventional model (Figure 2B). Thus it is likely that the decrease rate of *I* simulated from the conventional *SEIR* model after passing the peak may be slower than the decrease rate of actual daily cases.

An application of the new *SEIR* model in analysis of epidemic data in Excel is demonstrated in Figure 4. In this application, we simulated the number of daily new COVID-19 cases in the United States. To do so, we need to first determine or estimate the parameters *l* and *c*, the initial values *S*_0_, *E*_0_, *I*_0_ and *R*_0_, the starting date that COVID-19 began to transmit in the United States, and the coefficient *α* in Eqn. (2e) ^17,21^. Then we can use Eqns. (4a), (4c) and (2e) to calculate *S*_*n*_ and *I*_n_ respectively. By regulating the time-dependent transmission coefficient *β*_*n*_, we can fit the calculated number of daily COVID-19 cases *αI*_n_ to the reported 7-days average number of daily COVID-19 cases 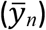 in the United States on a given day *n* after COVID-19 starts to spread in the United States. In this way, we determine *β*_*n*_ on each day until January 22, 2021. COVID-19 vaccines were given to people in the US were in the mid December 2020. In the above calculations, we did not consider the effect of COVID-19 vaccines on daily COVID-19 cases because the number of fully vaccinated people or even the number of people, who accepted at least one dose of COVID-19 vaccine, is much fewer than the number of susceptible people by January 22, 2021. It should be pointed out that derivation of Eqn. (4) needs an unstated condition: individuals who are recovered from the infectious disease will not be re-infected by this infectious disease again. Furthermore, the effect of vaccines on the infectious disease and breakthrough infections is not considered in the *SEIR* models. More complicated epidemic models considering effects of vaccines and breakthrough infections on the spread of infectious diseases are under study.

In conclusion, the underlying assumptions of the conventional *SIR* or *SIR*-derived models are not optimized for describing the transmission process of infectious diseases. A latent period-infectious period chronological order should be applied to these models for determining the relationship among the model variables. The new *SEIR* model based on the latent period-infectious period chronological order has improved the conventional models in simulating the transmission process of infectious diseases.

## Data Availability

All data produced in the present study are available upon reasonable request to the authors

## Notes

### Competing Interest Statement

The authors have declared no competing interest.

### Funding Statement

This study did not receive any funding

